# Emergency obstetric care access dynamics in Africa’s cities: Analysis of women’s self-reported care-seeking pathways in Kampala city, Uganda

**DOI:** 10.1101/2024.05.15.24307462

**Authors:** Birabwa Catherine, Beňová Lenka, van Olmen Josefien, Semaan Aline, Waiswa Peter, Banke-Thomas Aduragbemi

## Abstract

The rapid urbanization, particularly in Africa has posed several challenges that affect provision and accessibility of healthcare. The complex mix of providers, socio-economic inequalities, and inadequate infrastructure have been found to limit clients’ ability to reach and utilize routine and emergency health services. A growing body of literature shows poor health outcomes in African cities, including high institutional maternal mortality, which reflect poorly performing health systems. Understanding care-seeking pathways is necessary for improving health service delivery and ultimately improving health outcomes. We describe typologies and attributes of care-seeking pathways, using self-reported data from a cross-sectional survey of 433 women (15-49 years) who had obstetric complications, from nine health facilities in Kampala city. Participants’ average age was 26 (SD=6) years, and 55% (237/433) lived in the city suburbs. We identified four broad pathway sequences based on number and location of steps: pathways with one step, directly to a facility that could provide required care (42%, 183/433); two steps, mostly including direct referrals from basic and comprehensive obstetric facilities (40%, 171/433); three steps, including potentially delayed referral trajectories as women first return home (14%, 62/433); and ≥4 steps (4%, 17/433). Comprehensive obstetric facilities referred out 43% (79/184) of women who initially sought care in these facilities. Peripheral facilities referred 65% of women directly to the National Referral Hospital. Majority (60%, 34/57) of referred women first returned home before going to the final care facility. Our findings suggest that care pathways of women with obstetric complications in Kampala mostly comprise of at least two formal providers. This implies that efforts to strengthen urban health systems for maternal health should adopt broad or integrated approaches; and calls for improved inter-facility communication, streamlined referral processes and emergency transport availability. Future studies should investigate quality and experiences of care along the pathways.

**Key findings:**

- The pathways to care were multiple and varied by complication. Majority included at least two formal providers, giving insights into the dynamics surrounding obstetric referrals in the city such as time spent in a facility before referral and travel time between facilities.
- The issues identified from the facility referral pathways were: referrals from comprehensive obstetric facilities, bypassing of referral hierarchies by peripheral facilities, and low utilization of ambulances by peripheral facilities.
- Additionally, a notable percentage of referral pathway trajectories were interrupted and delays created therein, by women having to go back home after first seeking care from a formal provider. This was largely to obtain funds and other requirements.
- Majority of women seeking care for complications in Kampala city reside in neighboring suburbs.

**Key implications:**

- Policy makers and program stakeholders need to develop strategies and design services/systems that align with the care-seeking behaviors of clients in urban areas, including a substantial reliance on peripheral facilities, high contribution of referrals and the complexity of delays as clients move across different types or levels of providers.
- Policy makers should strengthen and expand coverage of comprehensive obstetric care to reduce unnecessary referrals or bypassing. Policy initiatives to facilitate access through private hospitals could contribute to improved access.
- Subnational stakeholders in urban areas, with support from national stakeholders, need to develop effective strategies and plans such as networks of care or care pathways for organizing and monitoring service delivery in metropolitan areas, with proper accountability for outcomes and resources.

## Introduction

Urbanization is a major ongoing trend worldwide, with approximately 55% of the world’s population living in urban areas in 2018.^1^ Urbanization holds potential for socio-economic development but is associated with several challenges such as urban poverty and increasing inequality, which disproportionately affect low- and middle-income countries.^2^ The urbanization process in Africa is more rapid than that in other continents, and is characterized by inequality, informal settlement expansion, and inadequate infrastructure, including healthcare.^3,4^ These features have implications for urban health systems and could exacerbate barriers to improving health outcomes in urban settings.^5,6^ Studies in African cities show poor maternal and perinatal health outcomes such as high institutional maternal mortality and inequitable coverage of maternal health services.^7,8^ These partly reflect poorly performing urban health systems, in the context of a rapidly growing population with dynamic daily and seasonal migration patterns amidst constrained healthcare infrastructure.

Improving health outcomes is contingent on provision of quality health services, which also influences population access to healthcare.^9^ Urban areas generally have a high availability and coverage of health services compared to rural areas, even for maternal health.^7,10^ However, accessibility of quality care is challenged by complexities of urban health systems and their environment. For example, the high number and mix of providers result in failure or delay to seek healthcare and bypassing of health facilities because people do not know appropriate facilities to choose.^11^ Concurrently, dominance of fee-based private providers implies that essential care is unaffordable for majority urban poor.^12^ Furthermore, poor housing structures (slums/informal settlements) and inadequate road infrastructure also affect timely access.^6^ Long travel times, poor road infrastructure, and cost of both transport and care affect access to and use of maternal health services in urban settings.^13,14^ Understanding care-seeking pathways in urban settings is necessary for improving health service delivery and ultimately improving health outcomes.

Over 60% of global maternal deaths occur in sub-Saharan Africa (SSA), largely due to treatable obstetric complications such as postpartum hemorrhage and hypertension in pregnancy.^15,16^ As such, timely access to appropriate emergency obstetric care (EmOC) is critical for maternal and perinatal survival.^17^ EmOC includes nine interventions recommended for managing common obstetric complications. Facilities with capacity to provide seven of the nine interventions are designated as basic EmOC (BEmOC) facilities. These interventions include administration of parenteral uterotonics, antibiotics and anticonvulsants/antihypertensives, manually removing retained placenta, removing retained products of conception, performing assisted vaginal delivery and basic neonatal resuscitation. In addition to these, facilities conducting caesarean deliveries and providing blood transfusion are designated as comprehensive EmOC (CEmOC) facilities.^18^ Furthermore, quality EmOC includes appropriate referral that is, making timely decisions to refer, using appropriate transportation, and appropriate information exchange between health facilities.^19^

Studies on healthcare seeking in urban settings in Africa show varied patterns such as bypassing of primary health facilities for routine care and preference of public health facilities for maternal health services.^13,20^ Regarding EmOC particularly, studies have focused on the determinants of access,^21^ with a few studies describing pathways (series of steps women take along their care-seeking journeys) to care.^22–24^ Studies in urban settings in SSA elaborate travel dynamics of women in emergency situations, highlighting different patterns and determinants.^22,25,26^ However, these insights emerge from a few cities in the region, yet women’s care-seeking processes might differ in other cities due to differences in resource availability, governance structures and population dynamics, which influence organization of and timely access to health services.^27^ Additionally, existing studies largely use data from medical records rather than women’s first-hand accounts and provide less information on intermediate steps between place of origin and where women receive care. Insights from women’s lived experiences provide an opportunity to integrate women’s voices in service delivery planning/strategies and improve health system responsiveness.

Uganda has made substantial progress in reducing maternal mortality,^28^ however, poor maternal and perinatal health outcomes in the country’s capital city (Kampala) remain a challenge.^29^ Persistent poor health outcomes amidst healthcare deprivations and a struggling healthcare system in Kampala present important research areas with potential relevance for other cities in Africa.^29,30^ In 2020, the Ugandan government prioritized urban areas with a strategic focus on improving the organization of health services and health system performance.^31^ Existing evidence in Kampala related to pathways to EmOC describes conditions for which women are referred between health facilities, and sources of obstetric referrals.^32–34^ However, this information is limited to a few public health facilities and there is insufficient evidence on the steps women actually take to reach care. This study’s objective was to examine and sequentially map self-reported care-seeking pathways among women who had obstetric complications in selected EmOC facilities in Kampala city, Uganda.

## Methods

### Study design and setting

This cross-sectional study was conducted in EmOC facilities in Kampala city, Uganda’s oldest and largest urban area, part of the Greater Kampala Metropolitan Area together with Wakiso, Mukono and Mpigi districts. Kampala has a resident population of 1.7 million which swells to 3.5 million during the day.^33^ Kampala exhibits economic disparities with poverty rates as high as 25% in some parishes.^35^ Kampala’s health system, comprised of a mix of 1,497 public and private health facilities in 2020,^33^ operates through different levels ranging from health centres (HCs) to National Referral Hospitals (NRH) (Figure 1). Uncomplicated deliveries can occur at any level, but obstetric complications are managed in CEmOC facilities (HC IV and above).^36,37^ Although privately-owned facilities are the majority (98%, n=1471), only 2% (n=30) are designated CEmOC facilities, and women predominantly seek obstetric care from the 26 (2%) public facilities which are expected to provide free care.^13^ We purposefully selected nine EmOC facilities, seven public and two private (Figure 1). We aimed to include typical providers of obstetric care across Kampala’s administrative divisions.^36,37^ Eight of the selected facilities are among those conducting the highest number of deliveries in Kampala.

**Figure. 1.**
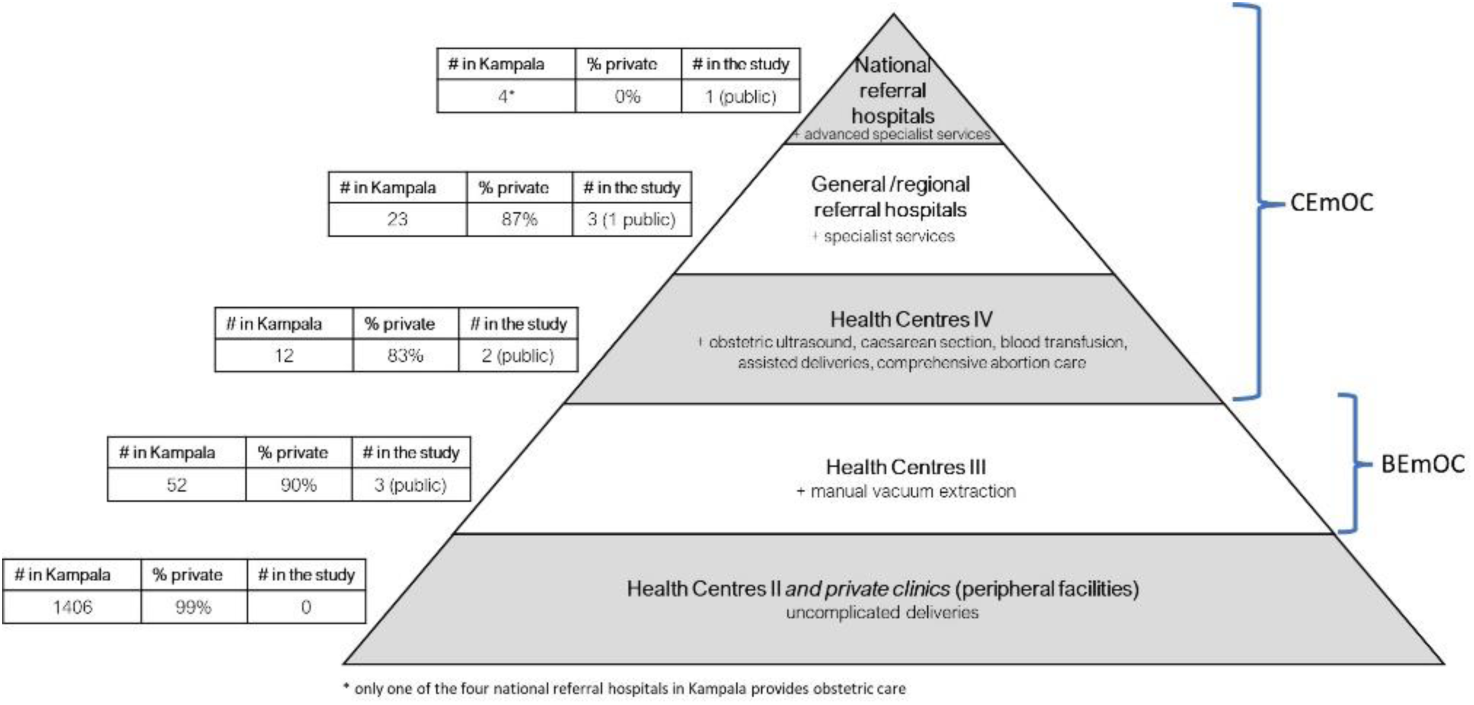
Health service delivery structure in Uganda for obstetric care and facilities included in the study. **Source**: developed for the study based on Uganda’s health service delivery standards^33,37^

### Study participants

Women who had obstetric complications were approached for inclusion in this study. We included all direct obstetric complications-obstetric hemorrhage (antepartum and postpartum), pre-eclampsia/eclampsia, obstructed/prolonged labor, pregnancy-related sepsis, complications of abortion and ectopic pregnancy. Women aged 15-49 years who provided written consent to participate in the study were included. Eligible women who declined to or could not participate were not included.

### Sampling and recruitment

Using a 50% prevalence for pathways to EmOC within which women are referred from one facility to another^34^ and a 10% adjustment for non-response, we calculated a sample size of 428 women. We recruited participants from different units/wards within the facilities, including maternity/labor, postnatal, high-dependency unit, gynecology, and emergency departments. We reviewed women’s medical files to identify those with an obstetric complication and confirmed this through consultation with unit/ward in-charge (a midwife or senior nursing officer) or doctors on duty. Participants were selected consecutively throughout the study period. We aimed to select at least a half of the sample size from the NRH due to referral patterns in Kampala^33^. To ensure variation across the six complications at the NRH, recruitment of obstructed labor cases was stopped as their admissions superseded other complications. In all, we approached 440 women, of which seven declined to participate.

### Study variables

Our primary outcome was self-reported care-seeking pathways, retrospectively captured by asking women to detail steps taken from deciding to seek help for symptoms or going to a health facility for childbirth or routine care, up to their final care facility. We utilized a sequential approach informed by Haenssgen and Ariana^38^ to capture each step’s location, activities, and duration. Locations included different health facilities, informal providers (like traditional birth attendants) and places of self-care/social support (home for the participant or another person). We captured any reported activity done or help received at each step. These included formal care (such as assessment, referral, delivery), self/informal care (including preparing childbirth items), and ignoring. We captured self-reported time spent in each step and between steps, initially categorized as minutes, hours, days, and weeks, and then quantified. Additional data on transportation, enablers and barriers faced were collected. Descriptions of pathway attributes and other study variables are provided in Table 1.

**Table 1:**
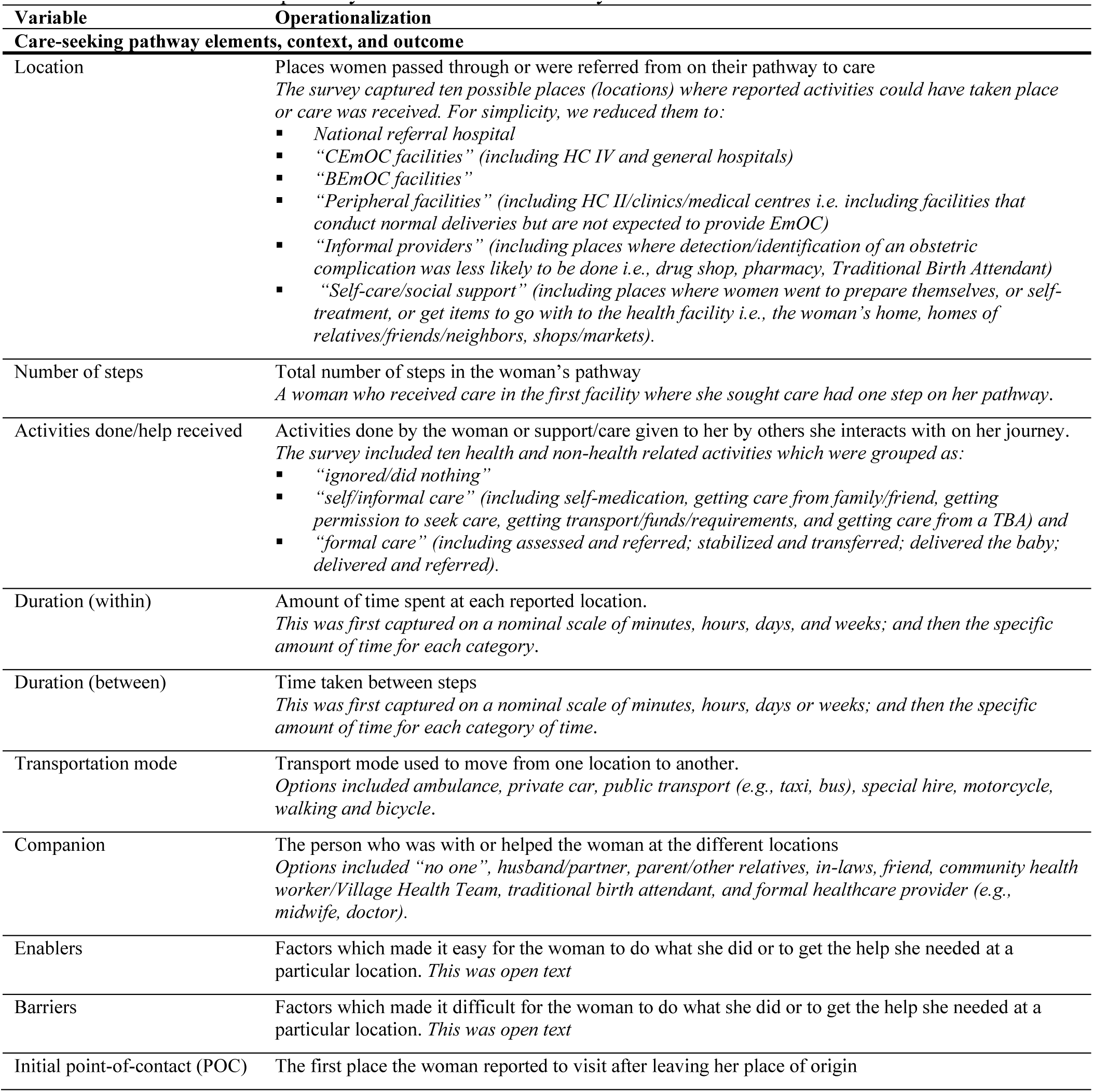
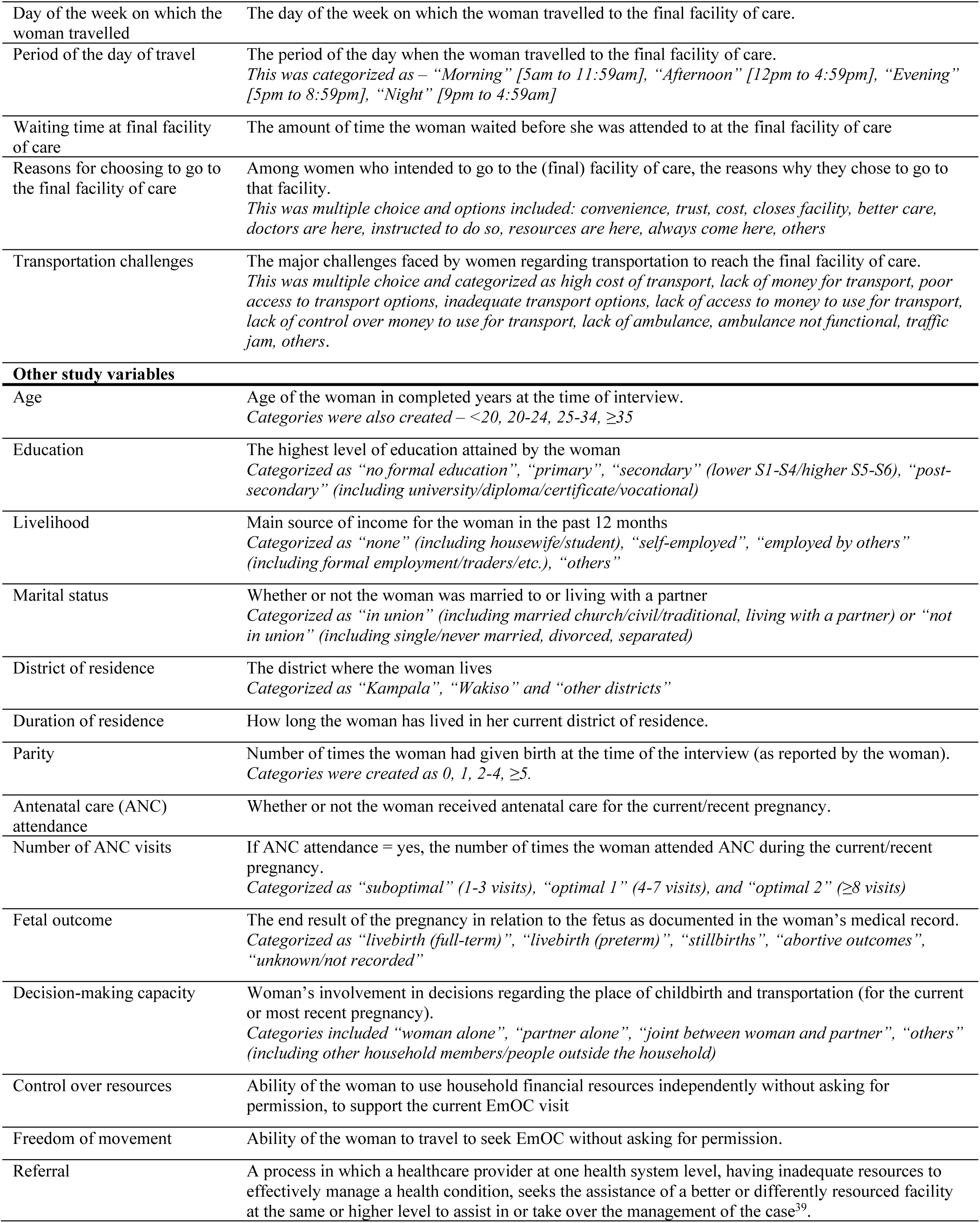
Definition of pathway attributes and other study variables.

| Variable | Operationalization |
| --- | --- |
| **Care-seeking pathway elements, context, and outcome** |  |
| Location | Places women passed through or were referred from on their pathway to care<br>*The survey captured ten possible places (locations) where reported activities could have taken place or care was received. For simplicity, we reduced them to:*<br>▪ National referral hospital<br>▪ “CEmOC facilities” (including HC IV and general hospitals)<br>▪ “BEmOC facilities”<br>▪ “Peripheral facilities” (including HC II/clinics/medical centres i.e. including facilities that conduct normal deliveries but are not expected to provide EmOC)<br>▪ “Informal providers” (including places where detection/identification of an obstetric complication was less likely to be done i.e., drug shop, pharmacy, Traditional Birth Attendant)<br>▪ “Self-care/social support” (including places where women went to prepare themselves, or self-treatment, or get items to go with to the health facility i.e., the woman’s home, homes of relatives/friends/neighbors, shops/markets). |
| Number of steps | Total number of steps in the woman’s pathway<br>*A woman who received care in the first facility where she sought care had one step on her pathway.* |
| Activities done/help received | Activities done by the woman or support/care given to her by others she interacts with on her journey.<br>*The survey included ten health and non-health related activities which were grouped as:*<br>▪ “ignored/did nothing”<br>▪ “self/informal care” (including self-medication, getting care from family/friend, getting permission to seek care, getting transport/funds/requirements, and getting care from a TBA) and<br>▪ “formal care” (including assessed and referred; stabilized and transferred; delivered the baby; delivered and referred). |
| Duration (within) | Amount of time spent at each reported location.<br>*This was first captured on a nominal scale of minutes, hours, days, and weeks; and then the specific amount of time for each category.* |
| Duration (between) | Time taken between steps<br>*This was first captured on a nominal scale of minutes, hours, days or weeks; and then the specific amount of time for each category of time.* |
| Transportation mode | Transport mode used to move from one location to another.<br>*Options included ambulance, private car, public transport (e.g., taxi, bus), special hire, motorcycle, walking and bicycle.* |
| Companion | The person who was with or helped the woman at the different locations<br>*Options included “no one”, husband/partner, parent/other relatives, in-laws, friend, community health worker/Village Health Team, traditional birth attendant, and formal healthcare provider (e.g., midwife, doctor).* |
| Enablers | Factors which made it easy for the woman to do what she did or to get the help she needed at a particular location. *This was open text* |
| Barriers | Factors which made it difficult for the woman to do what she did or to get the help she needed at a particular location. *This was open text* |
| Initial point-of-contact (POC) | The first place the woman reported to visit after leaving her place of origin |
| Day of the week on which the woman travelled | The day of the week on which the woman travelled to the final facility of care. |
| Period of the day of travel | The period of the day when the woman travelled to the final facility of care.<br>*This was categorized as – “Morning” [5am to 11:59am], “Afternoon” [12pm to 4:59pm], “Evening” [5pm to 8:59pm], “Night” [9pm to 4:59am]* |
| Waiting time at final facility of care | The amount of time the woman waited before she was attended to at the final facility of care |
| Reasons for choosing to go to the final facility of care | Among women who intended to go to the (final) facility of care, the reasons why they chose to go to that facility.<br>*This was multiple choice and options included: convenience, trust, cost, closes facility, better care, doctors are here, instructed to do so, resources are here, always come here, others* |
| Transportation challenges | The major challenges faced by women regarding transportation to reach the final facility of care.<br>*This was multiple choice and categorized as high cost of transport, lack of money for transport, poor access to transport options, inadequate transport options, lack of access to money to use for transport, lack of control over money to use for transport, lack of ambulance, ambulance not functional, traffic jam, others.* |
| **Other study variables** |  |
| Age | Age of the woman in completed years at the time of interview.<br>*Categories were also created – <20, 20-24, 25-34, ≥35* |
| Education | The highest level of education attained by the woman<br>*Categorized as “no formal education”, “primary”, “secondary” (lower S1-S4/higher S5-S6), “post-secondary” (including university/diploma/certificate/vocational)* |
| Livelihood | Main source of income for the woman in the past 12 months<br>*Categorized as “none” (including housewife/student), “self-employed”, “employed by others” (including formal employment/traders/etc.), “others”* |
| Marital status | Whether or not the woman was married to or living with a partner<br>*Categorized as “in union” (including married church/civil/traditional, living with a partner) or “not in union” (including single/never married, divorced, separated)* |
| District of residence | The district where the woman lives<br>*Categorized as “Kampala”, “Wakiso” and “other districts”* |
| Duration of residence | How long the woman has lived in her current district of residence. |
| Parity | Number of times the woman had given birth at the time of the interview (as reported by the woman).<br>*Categories were created as 0, 1, 2-4, ≥5.* |
| Antenatal care (ANC) attendance | Whether or not the woman received antenatal care for the current/recent pregnancy. |
| Number of ANC visits | If ANC attendance = yes, the number of times the woman attended ANC during the current/recent pregnancy.<br>*Categorized as “suboptimal” (1-3 visits), “optimal 1” (4-7 visits), and “optimal 2” (≥8 visits)* |
| Fetal outcome | The end result of the pregnancy in relation to the fetus as documented in the woman’s medical record.<br>*Categorized as “livebirth (full-term)”, “livebirth (preterm)”, “stillbirths”, “abortive outcomes”, “unknown/not recorded”* |
| Decision-making capacity | Woman’s involvement in decisions regarding the place of childbirth and transportation (for the current or most recent pregnancy).<br>*Categories included “woman alone”, “partner alone”, “joint between woman and partner”, “others” (including other household members/people outside the household)* |
| Control over resources | Ability of the woman to use household financial resources independently without asking for permission, to support the current EmOC visit |
| Freedom of movement | Ability of the woman to travel to seek EmOC without asking for permission. |
| Referral | A process in which a healthcare provider at one health system level, having inadequate resources to effectively manage a health condition, seeks the assistance of a better or differently resourced facility at the same or higher level to assist in or take over the management of the case<sup>39</sup>. |

### Data collection

Data were collected using an interviewer-administered questionnaire developed for the study, using KoboCollect. An excerpt of the questions used to generate the pathway data is provided in (Appendix Figure. 1). Nine research assistants, trained and supervised by the lead author, collected the data. These included midwives and graduates with prior experience in facility-based research. We reviewed women’s medical records for information on diagnosis on admission, pregnancy outcome and gestation age. Data were collected between July and September 2022.

### Data analysis

Data were cleaned and analyzed in Stata v14 (Stata Corporation, College Station, Texas, US). Descriptive statistics including frequencies and percentages were used to summarize categorical characteristics of participants and pathway attributes; the mean and standard deviation (SD) were used to summarize age across the sample, while the median and inter-quartile range (IQR) were calculated for duration (within/between steps). Data were presented in tables and figures.

We analyzed all reported pathways to identify unique sequences, and then generated a typology of common pathways using the number of steps and their location. At each step, we mapped engagement with different sources of care/help along the pathways by counting each location. For each common pathway sequence, we summarized relevant attributes, considering key obstetric care practices and the urban context. During analysis of pathways involving facility referral, we distinguished CEmOC from other health facilities. We described individual pathways of selected women (using pseudonyms) to illustrate common pathway sequences. Next, we explored pathways by complication, analyzing the distribution of common pathways, travel and referral patterns, and how pathways started. We considered two possible ways in which women’s care-seeking started: 1) symptoms (a woman experienced something which prompted her to seek care – *woman sensed something could be wrong*); and 2) labor (A woman went to the facility for routine childbirth care and in the process of or after birth developed the complication – *woman was not aware something could be wrong with the pregnancy when she first sought care*). Instances where a woman’s journey started going to a health facility for routine care such as ANC and the complication detected during this time (signs), were summarized with symptoms.

### Ethics

Ethical approval was obtained from the ethics committees of Makerere University School of Public Health (SPH-2021-169), the Uganda National Council for Science and Technology (HS1952ES), the Institute of Tropical Medicine Antwerp (1529/21) and the University of Antwerp Hospital (2021-1743). Written consent to participate in the study was obtained from each participant before the interview. No directly identifying information such as individual names and national identification numbers were collected. Each participant and study site were given a unique identifier.

## Results

### Participant characteristics

Among the 433 participants, 137 (32%) had obstructed labor, 105 (24%) had pre-eclampsia/eclampsia, 87 (20%) had hemorrhage, 54 (13%) had ectopic pregnancy complications, 50 (12%) had abortion-related complications and 26 (6%) had pregnancy-related sepsis. Women in the sample were 26 years on average (SD=6). The majority were married/cohabiting (79%), had secondary education (53%), and lived outside Kampala city (55%) particularly in Wakiso district (Table 2). More than 70% attended ANC, with a maximum of nine visits. Among all participants, 5% experienced a stillbirth. Place of delivery was more commonly decided by women alone (34%), while decisions related to transportation (33%) and spending money for pregnancy/childbirth care (39%) were more frequently made by women’s partners. Further, 32% of participants first sought permission to spend money to seek care, and this was mostly from their partners (89%, 117/131).

**Table 2.** Characteristics of the sample of women who experienced obstetric complications in Kampala (n=433, 9 facilities)

| Characteristic | Categories | Frequency (n) | Percentage (%) |
| --- | --- | --- | --- |
| Age group | <20 years | 49 | 11.3 |
|  | 20-24 years | 146 | 33.7 |
|  | 25-34 years | 189 | 43.7 |
|  | ≥35 years | 49 | 11.3 |
|  | Mean: 26.0, SD=6 |  |  |
| Marital status (n=431) | Single/divorced/separated | 89 | 20.7 |
|  | Married/cohabiting | 342 | 79.3 |
| Education level (n=432) | None/never gone to school | 5 | 1.2 |
|  | Primary | 105 | 24.3 |
|  | Secondary | 227 | 52.5 |
|  | Post-secondary | 95 | 22.0 |
| Livelihood (n=430) | None (e.g., housewife)/student | 155 | 36.0 |
|  | Self-employed | 105 | 24.4 |
|  | Employed by others | 146 | 34.0 |
|  | Others | 24 | 5.6 |
| District of residence | Kampala | 196 | 45.2 |
|  | Wakiso | 199 | 46.0 |
|  | Other districts | 38 | 8.8 |
| Duration of residence | <1 month | 4 | 0.9 |
|  | Months | 90 | 20.8 |
|  | Years | 315 | 72.8 |
|  | Always | 24 | 5.5 |
| ANC attendance | No | 98 | 22.6 |
|  | Yes | 335 | 77.4 |
| Number of ANC visits (n=335) | 1-3 visits (sub-optimal) | 91 | 27.1 |
|  | 4-7 visits (optimal 1) | 220 | 65.7 |
|  | 8-9 visits (optimal 2) | 10 | 3.0 |
|  | No response/don't know | 14 | 4.2 |
| Parity | 0 | 30 | 6.9 |
|  | 1 | 173 | 40.0 |
|  | 2-4 | 194 | 44.3 |
|  | ≥5 | 38 | 8.8 |
| Fetal outcome | Livebirth (full-term) | 223 | 51.5 |
|  | Livebirth (preterm) | 73 | 16.9 |
|  | Stillbirth | 21 | 4.8 |
|  | Abortive outcome | 101 | 23.3 |
|  | Not recorded | 15 | 3.5 |
| Decision on place of delivery | Woman alone | 145 | 33.5 |
|  | Joint decision† | 115 | 26.6 |
|  | Partner alone | 100 | 23.1 |
|  | Others* | 59 | 13.6 |
|  | No response | 14 | 3.2 |
| Decision on transportation to the health facility | Woman alone | 116 | 26.8 |
|  | Joint decision† | 102 | 23.5 |
|  | Partner alone | 141 | 32.6 |
|  | Others* | 70 | 16.2 |
|  | No response | 4 | 0.9 |
| Decision on how to spend money for pregnancy/childbirth care | Woman alone | 91 | 21.1 |
|  | Joint decision† | 124 | 28.6 |
|  | Partner alone | 169 | 39.0 |
|  | Others* | 45 | 10.4 |
|  | No response | 4 | 0.9 |
| Had to request permission to spend money for pregnancy/childbirth care from another person | No | 296 | 68.4 |
|  | Yes | 137 | 31.6 |
| Had to request permission to go to the facility | No | 292 | 67.4 |
|  | Yes | 141 | 32.6 |

†Joint between the woman and her partner
\*'Others' includes other household members, people outside the household, joint decision between woman and other household members.

### Pathways to care among women who had obstetric complications

We identified 51 unique pathways among the 433 participants (Appendix Figure 2). Across all pathways, the initial points-of-contact were as follows: CEmOC facilities (43%, 184/433), peripheral facilities (25%, 110/433), NRH (14%, 60/433), BEmOC facility (12%, 50/433), self-care/social support (5%, 20/433) and informal care (2%, 9/433). Of the 184 participants who first sought care from a CEmOC facility, 43% (n=79) were referred out. We grouped the pathways into four sequences (A, B, C, D) based on the total number (ranging from one to six) and location of steps taken (Figure 2). **Sequence A,** with women who had one step on the pathway (direct to the final facility of care), was the most common (42%, 183/433). The most frequent complication in this sequence was obstructed labor (36%, 65/183). **Sequence B** represents pathways with two steps including the final facility of care (40%, 171/433). The first step was a CEmOC facility (*seq. B1*) in 40% (69/171), a peripheral facility (*seq. B2*) in 39% (66/171), a BEmOC facility (*seq. B3*) in 17% (29/163), and a place of self-care/social support in 4% (7/171, not shown). The most frequent complication in sequence B was pre-eclampsia (32%, 55/171). **Sequence C** contains pathways with three steps including the final facility of care and accounted for 14% (62/433) of the sample. The most frequent complication in this sequence was pre-eclampsia (29%, 18/62). **Sequence D** represents pathways with four or more steps including the final facility of care, representing 4% (17/433) of the sample, with obstructed labor being the most common complication (41%, 7/17). Figure 2 also shows distribution of places women visited at each location (step) of the care pathways. Women who did not go directly to final facility (n=250) more commonly first sought care from peripheral (44%, n=109) and CEmOC (32%, n=79) facilities. At subsequent steps, places of self-care/social were the most accessed (e.g., 58% at step 2; 46/79).

**Figure 2.**
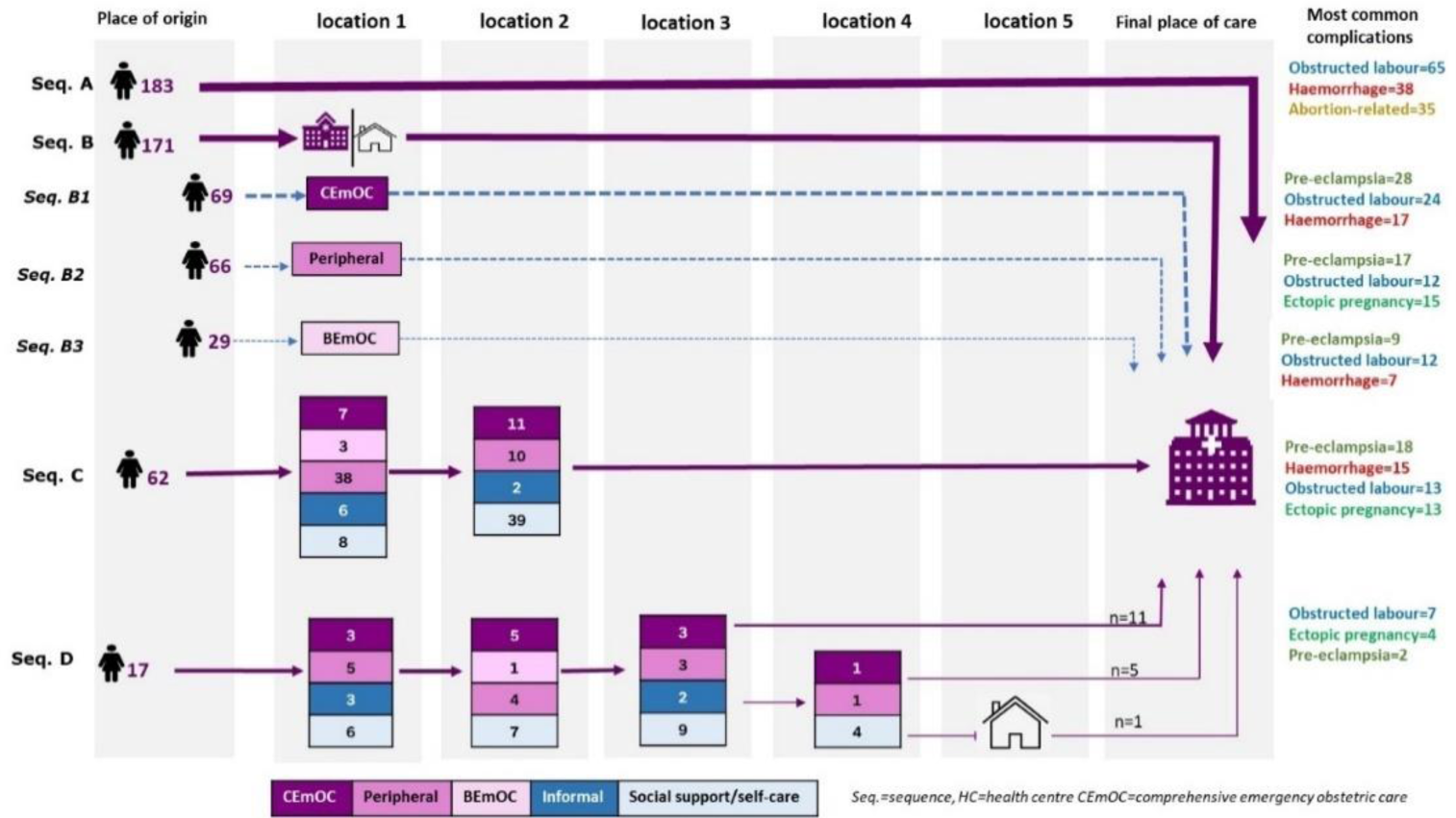
Common pathways to emergency obstetric care among women from selected facilities in Kampala city, n=433. Continuous arrows between the place of origin and ‘location 1’ in the figure show the major pathway sequences and the dashed arrows show sub-sequences. 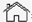 = home

### Description of selected attributes of the common pathway sequences to EmOC

Women with Sequence A (direct to final care facility) travelled mostly (54%) by motorcycle, with majority going to HC IVs and hospitals (Table 3). Having someone to help women prepare was an important facilitator for care-seeking in this group. Among participants with Sequence B pathways, particularly those involving health facilities (sub-Sequences B1-B3), transport availability facilitated care access for at least 20% of participants. The median time spent in the facility before referral was 4 hours (IQR=1-33) in CEmOC, 5 hours (IQR=1-14) in BEmOC and 2 hours (IQR=1-7) in peripheral facilities. Fifty-five percent of women referred by CEmOC facilities and 20% of women referred by BEmOC or peripheral facilities used an ambulance for transport to the final facility. Among participants with Sequence C, pathways of 24% of women included two formal health facilities before the final facility. Also, places of self-care/social support accounted for 60% as the second step after women first seeking care from a health facility. All 17 women with sequence D had unique pathway sequences, with six including more than one formal health facility before the final facility. Additional details on each sequence are provided in (Appendix Table 1).

**Table 3:** Key attributes of the major pathways among women who experienced obstetric complications in selected facilities in Kampala (n=433†)

| Pathway sequence | Most common transport mode to the final facility of care | Median travel time to the final facility | Main transport challenge(s)* | % going to NRH | Main enabler(s) | District of residence | Other notes |
| --- | --- | --- | --- | --- | --- | --- | --- |
| Direct pathways (Sequence A, n=183)<br>**Fig. 3.1** | ▪ Motorcycle 54% (99/182) | 0.5 hours (IQR=0.5-1) | ▪ Financial constraints (51%, 45/88)<br>▪ Traffic jam (44%, 39/88)<br>▪ Inadequate transport options (31%, 27/88) | 33% (60/183) | • Social support from partners, relatives and friends (51%, 93/183) | Kampala 53% (97/183) | Reasons for choosing the facility<br>▪ Better care (53%, 88/166)<br>▪ Convenient access (31%, 51/166)<br>▪ Usual place of care (26%, 43/166).<br>57%(n=105) went to HC IVs and hospitals; and 10%(n=18) went to BEmOC facilities |
| Pathways with two steps (Sequence B1, n=69)-<br>*Referral by CEmOC facilities*<br>**Fig. 3.2** | ▪ Ambulance 55% (38/69)<br>▪ Motorcycle 32% (22/69) | 1 hour (IQR=0.5-1.5). | ▪ Financial constraints (71%,27/38)<br>▪ Traffic jam (40%, 15/38)<br>▪ Inadequate transport options (26%,10/38) | 82%, 65/79 | • Transport availability (38%, 26/69) | Outside Kampala 65% (45/69) | ▪ Nearly 70% of women of women who had pre-eclampsia (31/46) and first sought care a CEmOC facility were referred out.<br>▪ Women spent a median of 4 hours (IQR=1-33) in the facility before being referred. |
| Pathways with two steps (Sequences B2 and B3, n=95)- *Referral by BEmOC or peripheral facilities*<br>**Fig. 3.3** | ▪ Motorcycle 39% (37/95)<br>▪ Ambulance 20% (19/95) | 1 hour (IQR=0.5-1) | ▪ Traffic jam (46%, 24/52)<br>▪ Inadequate transport options (42%,22/52)<br>▪ Financial constraints (39%,20/52) | 65% (62/95) | ▪ Transport availability 25% (24/95) | Outside Kampala 62% (59/95) | ▪ 67% (95/142) of women who first sought care at BEmOC or peripheral facilities went directly to the final facility of care.<br>▪ 48% (46/95) had gone for a check-up because they were not feeling well; while 40% (38/95) had gone to give birth<br>▪ Majority had ectopic pregnancy complications (52%, 28/54)<br>▪ Women spent a median of 3 hours (IQR=1-8) before being referred; longer in BEmOC (5hours; IQR=1-14) than in peripheral (2hours; IQR=1-7) facilities |
| Pathways with three steps (Sequence C, n=62)<br>**Fig. 3.4** | ▪ The first place visited was a peripheral facility in 61% (38/62)<br>▪ The last place visited before women reached the final facility of care was a place of self-care/social support in 63% (39/62)<br>▪ 24% (15/62) of women had pathways that included two formal providers before the final facility<br>○ Between two CEmOC facilities (2/15)<br>○ From peripheral and CEmOC facilities (7/15)<br>○ Between two peripheral facilities (6/15)<br>▪ Among women with pathways where the first step was a formal provider, places of self-care/social support accounted for 60% (34/57) of places visited at the second step.<br>○ This was highest in pathways of women treated for pre-eclampsia/eclampsia (71%, 12/17) and obstructed labor (70%, 7/10).<br>○ The most common activity done among the 34 women was obtaining childbirth requirements including money (65% 22/34).<br>▪ Majority lived outside Kampala 53% (33/62)<br>▪ 54% (34/62) ended at the NRH |  |  |  |  |  |  |
| Pathways with four or more steps (Sequence D, n=17)<br>**Fig. 3.5** | ▪ All 17 pathways for sequence D were unique.<br>▪ 35% (6/17) of women had pathways that included more than one formal provider before reaching the final facility of care.<br>▪ Majority lived outside Kampala 65% (11/17).<br>▪ 23% (4/17) ended at the NRH. |  |  |  |  |  |  |

NRH=National Referral Hospital.
\*Computed among women who reported to have experienced challenges with transportation to the facility; multiple answers possible so totals exceed 100%
†Some missingness in some denominators due to missing answers in the survey

### Individual participant pathways

Illustrations of women’s individual journeys in each sequence described in Table 3 are shown in Figure 3. Starting from the place of origin, the figures show how women with different complication, age and obstetric history profiles reached the final facility, and their outcomes. Figures 3.1 and 3.5 illustrate the variation in pathways women with the same complication can take to care.

**Figure 3.**
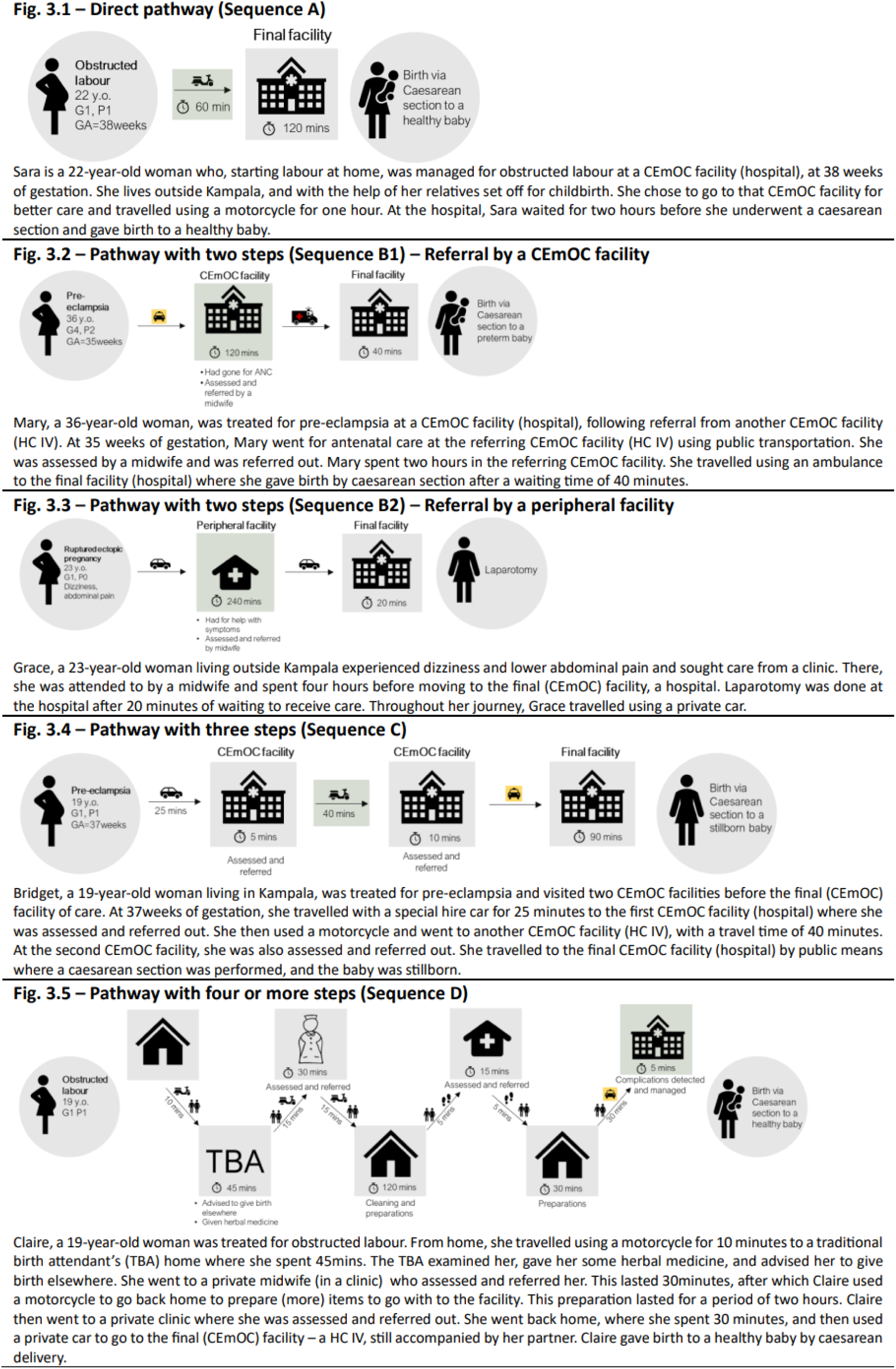
Individual pathways illustrating the pathway sequences. All women’s names are pseudonyms

### Description of pathways by obstetric complication

Pathways among women with sepsis (73%, 19/26), abortion (100%, 50/50), and ectopic pregnancy complications (98%, 53/54) mostly started with symptoms recognized by the woman or signs detected by a health worker (Appendix Figure 3). Among women with hemorrhage (63%, 55/87) and obstructed labor (91%, 125/137), pathways mostly started with women seeking care expecting routine childbirth (labor). Pathways of women with abortion complications mostly had one step (70%, 35/50), while over 50% pathways of women with all other complications had two or more steps. Additionally, women more frequently started their journey to the final facility of care in the morning across the six complications (168/428, 39%); others travelled in the afternoon (121/428, 28%), evening (68/428, 16%) and night (71/428, 17%). Over 50% of women across all complications, had pathways where the first facility visited was a designated CEmOC facility, except for ectopic pregnancy complications, for which the majority of women first sought care from a BEmOC or peripheral facility (52%, 28/54).

## Discussion

Our study presents access dynamics for EmOC in African cities through examining care-seeking pathways in Kampala city. We found four common pathway sequences based on the number and nature of steps taken by women with complications: 1) direct pathways (42%), 2) pathways with two steps (40%), 3) pathways with three steps (14%), and 4) pathways with four or more steps (4%). Our results further showed that 40% of women who first sought care from CEmOC facilities were referred out. Among women who first sought care in a BEmOC or peripheral facility, 65% were referred directly to the NRH, thus bypassing expected levels of referral hierarchy. There was lower utilization of ambulances for transportation during referral from BEmOC or peripheral facilities (20%) compared to referrals out of CEmOC facilities (55%). Also, there were delays within referral sections of women’s pathways with the majority (60%) returning home before going to the final facility.

We found that four in 10 women took a direct path to care. This finding could be attributed to perceived quality of care and physical accessibility,^40^ which in our study influenced women’s health facility choice, in addition to familiarity with the health facility. Additionally, “*early referral*” advice given to women with high-risk pregnancies during ANC as is recommended in Uganda,^36^ could explain direct care-seeking. Our study showed that women’s social networks played a key role in enabling direct care-seeking, suggesting the importance of involving women’s network members in interventions addressing delays in care-seeking. A special focus on vulnerable women, lacking social support/capital for childbirth in urban areas is necessary.

Among pathways with two steps, our results showed that 40% of women initially seeking care from CEmOC facilities (predominantly HCIVs) were referred out, notably so among women who had pre-eclampsia (70%). This is inconsistent with the expected functionality of CEmOC facilities which should manage nearly all complications. Moreover, we found that 65% of women initially seeking care from BEmOC or peripheral facilities were referred directly to the NRH, showing non-adherence to referral guidelines.^36^ These suboptimal referral practices could be attributed to health system constraints such as a lack of skilled/critical human resources, commodities, and inadequate capacity of some CEmOC facilities to manage critically sick patients; or the inability of women to afford care in private hospitals and weak enforcement of referral guidelines.^41,42^ Furthermore, we found that the length of time women spent before being referred out ranged from 2 hours (in peripheral facilities) to 5 hours (in BEmOC facilities). While this may include pre-referral treatment or waiting for transportation, it could also indicate failure of providers to make timely decisions and unnecessarily delaying referrals. Capacity strengthening of health facilities in line with their designated levels of functionality and better care coordination are essential, and could reduce delays from unnecessary referral and bypassing, and also crowding at receiving facilities.

We found that fewer women referred out of BEmOC or peripheral facilities (20%) used an ambulance compared to women referred out of CEmOC facilities (55%). This indicates that peripheral facilities, which are expected to refer all major obstetric complications, have limited availability or accessibility to ambulance services. Kampala has a centralized coordination system for ambulances that targets public and private health facilities.^33,43^ Failure of BEmOC and peripheral facilities to use ambulances can arise when multiple requests are made, and priority is given to other types of facilities or patients. Additionally, some peripheral facilities may not be aware of available emergency transportation services and may have limited support to access them. These and other factors related to functionality of ambulances and administrative procedures may explain the apparent low use of ambulances by peripheral facilities. This may translate as additional costs incurred by women as they shoulder the burden of arranging and paying for transportation, and exacerbates inequalities in access to EmOC. Improving coverage of affordable ambulance services at peripheral facilities and in communities is crucial to address this issue.

We further found that 20% of women visited more than one place before reaching the final care facility (Sequences C and D), with nearly a third seeking care from two or more formal health facilities. This reflects complexity of care-seeking in urban areas, and may be influenced by the type/capacity of health facilities visited by women.^23^ This finding could further reflect deficiencies in health facilities’ capacity to manage obstetric complications. However, it could also be attributed to women’s preferences, perceived severity of their condition, or unacceptability and un-affordability of the facility to which they were referred.^40,44^

Surprisingly, we found that 60% of women referred from one facility to another returned home before going to the final care facility. This was common among women with pre-eclampsia (71%) and obstructed labor (70%). This pattern points to inadequate birth preparedness and complication readiness, as women reported that they had to first prepare themselves, get baby clothes or money during this time. Poor birth preparedness practices among urban residents have also been reported in Nigeria.^45^ This is likely a result of financial constraints and limited decision-making power of women regarding money for pregnancy/childbirth which would force women to delay care seeking.^13^ It could also be attributed to the system inadequately supporting pregnant women with childbirth essentials during their stay in health facilities, or a lack of national health insurance, as is the case in Uganda. Inevitably, women bear this burden which potentially delays their care-seeking.

We found that the majority of women seeking care for complications in Kampala reside in neighboring districts or further away. Several factors contribute to this, such as inadequate numbers or readiness of EmOC facilities in neighboring districts, or proximity to Kampala which facilitates physical access to facilities. Critically, we note the lack of a gatekeeping practice in Uganda’s health system (one can go wherever they want). While districts in Uganda are mandated to manage their population’s healthcare under the decentralization policy, there is limited guidance on collaboration across district health systems which is crucial for the metropolitan context of Kampala. Among ongoing efforts to address urbanization challenges in Uganda is the development of an Urban Health Strategy. It is critical that this strategy includes a system to coordinate service delivery including referral care in metropolitan areas.

This could leverage the networks of care approach where health facilities in the metropolitan area are purposefully linked, to optimize patient experiences and outcomes.^46^

### Strengths and limitations

With a large sample of 433 women who had common six obstetric complications, our study provides useful insights into different features of pathways to EmOC in an urban setting, which can be used to improve the provision of EmOC services and optimize health outcomes in urban health facilities. We included eight of the high obstetric volume facilities in Kampala. However, the study also had some limitations. First, due to recruiting from health facilities, the pathways identified may not represent journeys of women who fail to reach skilled routine or emergency obstetric care. However, given the high facility delivery rate of 94% in Kampala ^47^, the identified pathways provide a good reflection of how women possibly reach care. Also, our recruitment sites included only two private hospitals which might not be representative of all women seeking care in the several private health facilities across the city. Second, we used self-reported data which could have been affected by response bias. Third, we did not include women who experienced an emergency due to fetal complications and other maternal complications. Similarly, we did not include pathways of women who experienced maternal deaths and some women, particularly those who experienced poor pregnancy outcomes such as stillbirths, declined to participate in the study.

### Implications for future research

Our study focused on care-seeking processes of women with complications. Future studies should examine processes of care provision at the health facilities where women sought care to further understand the alignment between care-seeking and service delivery. Also, future studies could use objective measures for some pathway attributes such as time spent within and between different steps and integrate quality, experience, and cost of care assessment along women’s pathways as well as women’s preferences. We also suggest that studies including women who experienced poor perinatal outcomes be conducted to map their care-seeking processes and establish pathway attributes which are more likely to contribute to poor outcomes.

## Conclusions

We found four common pathway sequences to care in Kampala, with most women going directly to the final care facility. The majority of women referred between health facilities lived in neighboring districts, and bypassing was common. Also, most referred women delayed reaching final facility as they had to first prepare themselves. Increasing availability and readiness of CEmOC facilities is crucial to reduce CEmOC-to-CEmOC referrals or bypassing of nearest CEmOC facilities by lower-level facilities. Referral care and healthcare seeking competency in Kampala and surrounding districts should be improved at all system levels to achieve effective patient care pathways. The “networks of care” approach could be adopted, which would need capacity building of peripheral facilities, streamlining and strengthening the quality of facility linkages, and increasing availability of affordable emergency transport options, especially for emergencies originating in peripheral facilities. The development of a national health insurance system that covers the urban poor could greatly improve timely access to care. Future studies should investigate technical quality and experience of care received in these pathways. Last, women’s social context should be integrated in interventions addressing delays in seeking/reaching EmOC.

## Data Availability

All data produced in the present study are available upon reasonable request to the authors.

## Author contributions

CB conceptualized the study, analyzed the data and produced initial drafts of the paper. ABT, JVO, LB and PW reviewed the design and provided overall study supervision. LB and AS participated in the analysis and visualization of results. All authors reviewed and edited the initial drafts and approved the final version that was submitted.

## Acknowledgements

We thank the study participants and research assistants. We thank Dr Dinah Amongin for reviewing and providing feedback on the manuscript.

## Funding statement

CB was funded by the Belgian Directorate for Development Co-operation PhD sandwich programme. The funders had no role in study design, data collection and analysis, decision to publish, or preparation of the manuscript.

## Competing interests

Authors declare no competing interests.

## Appendix

**Figure 1.**
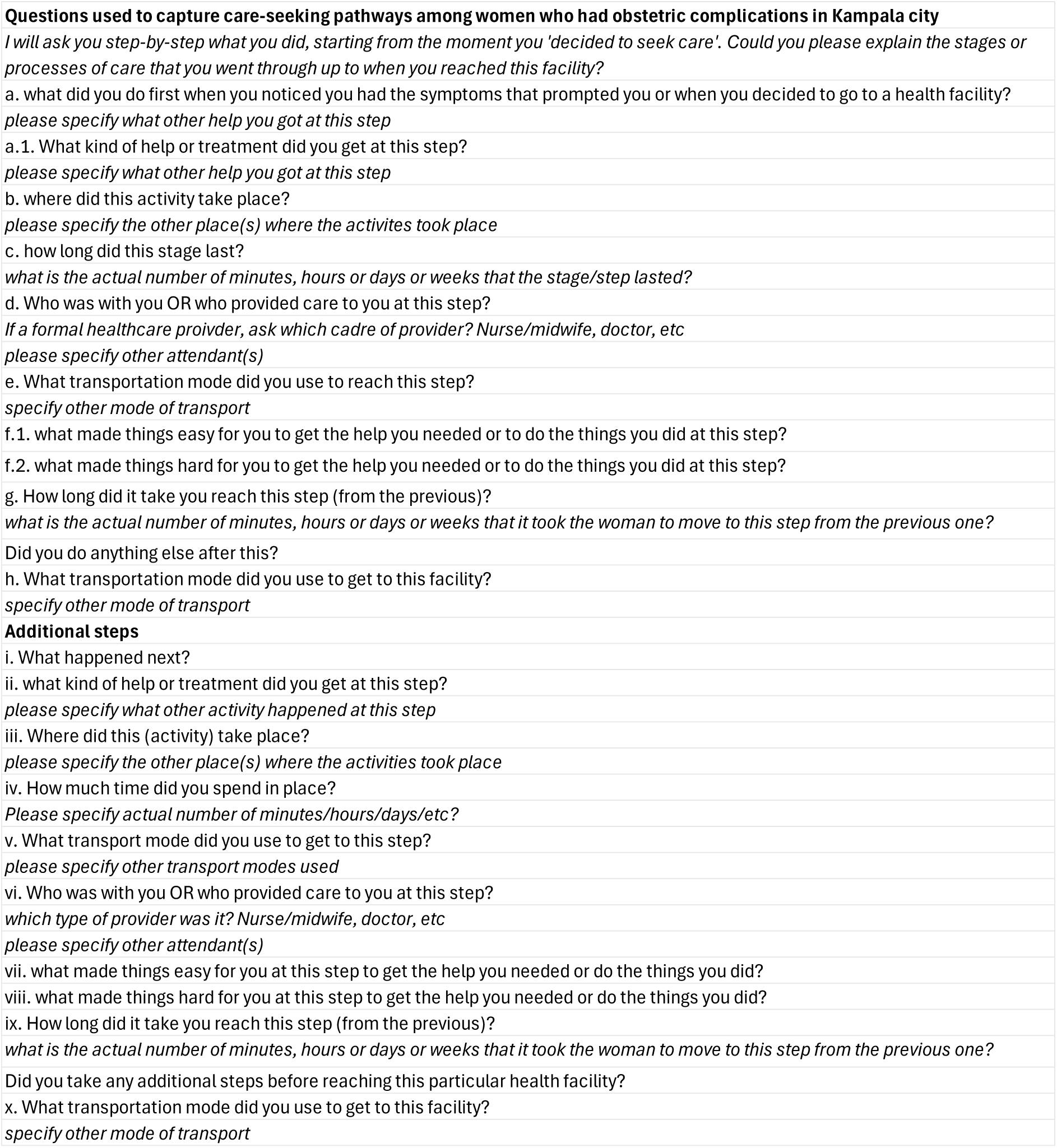
Questionnaire excerpt used to capture care-seeking pathways.

**Figure 2.**
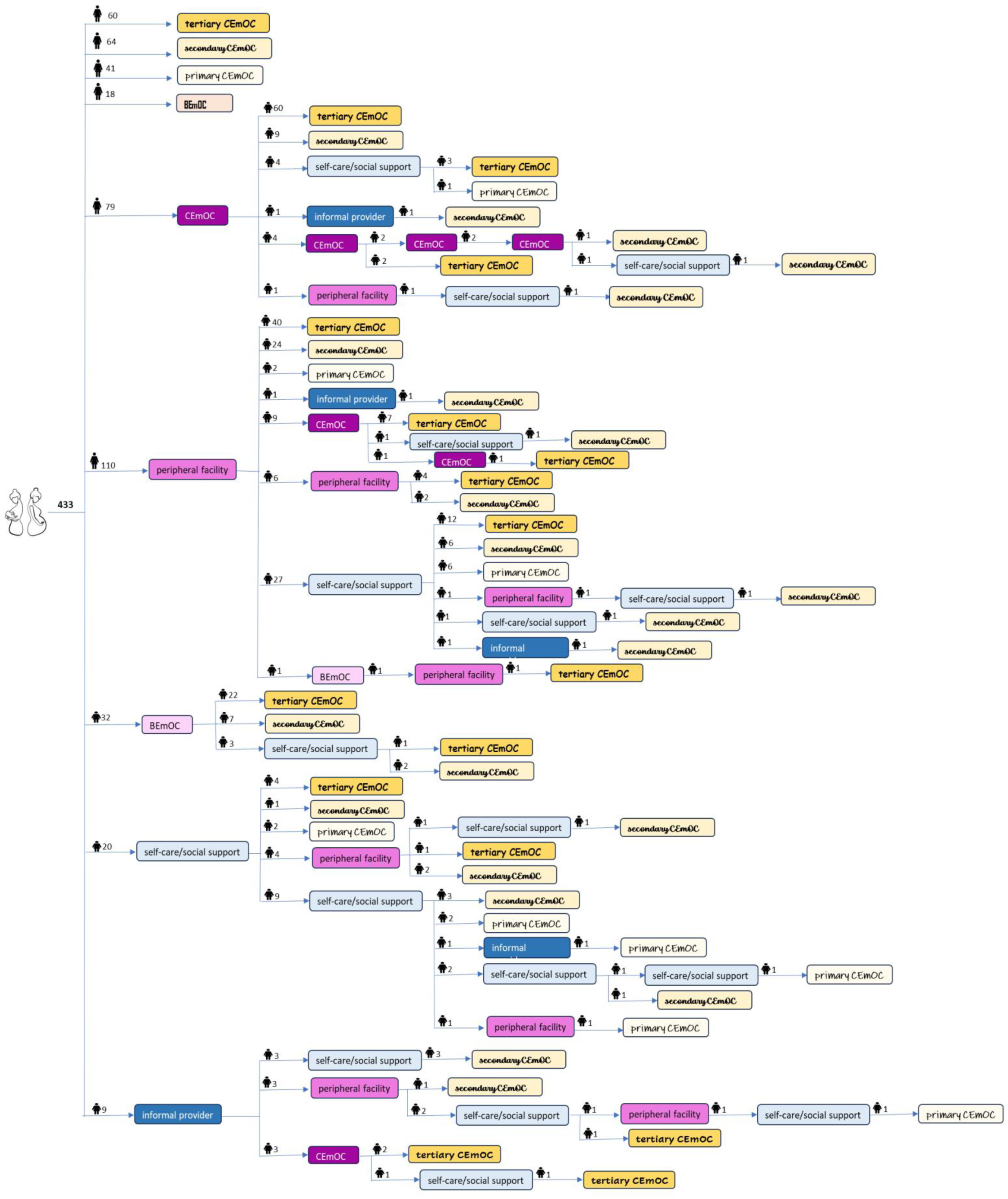
Unique pathway sequences identified across all 433 respondents.

**Figure 3.**
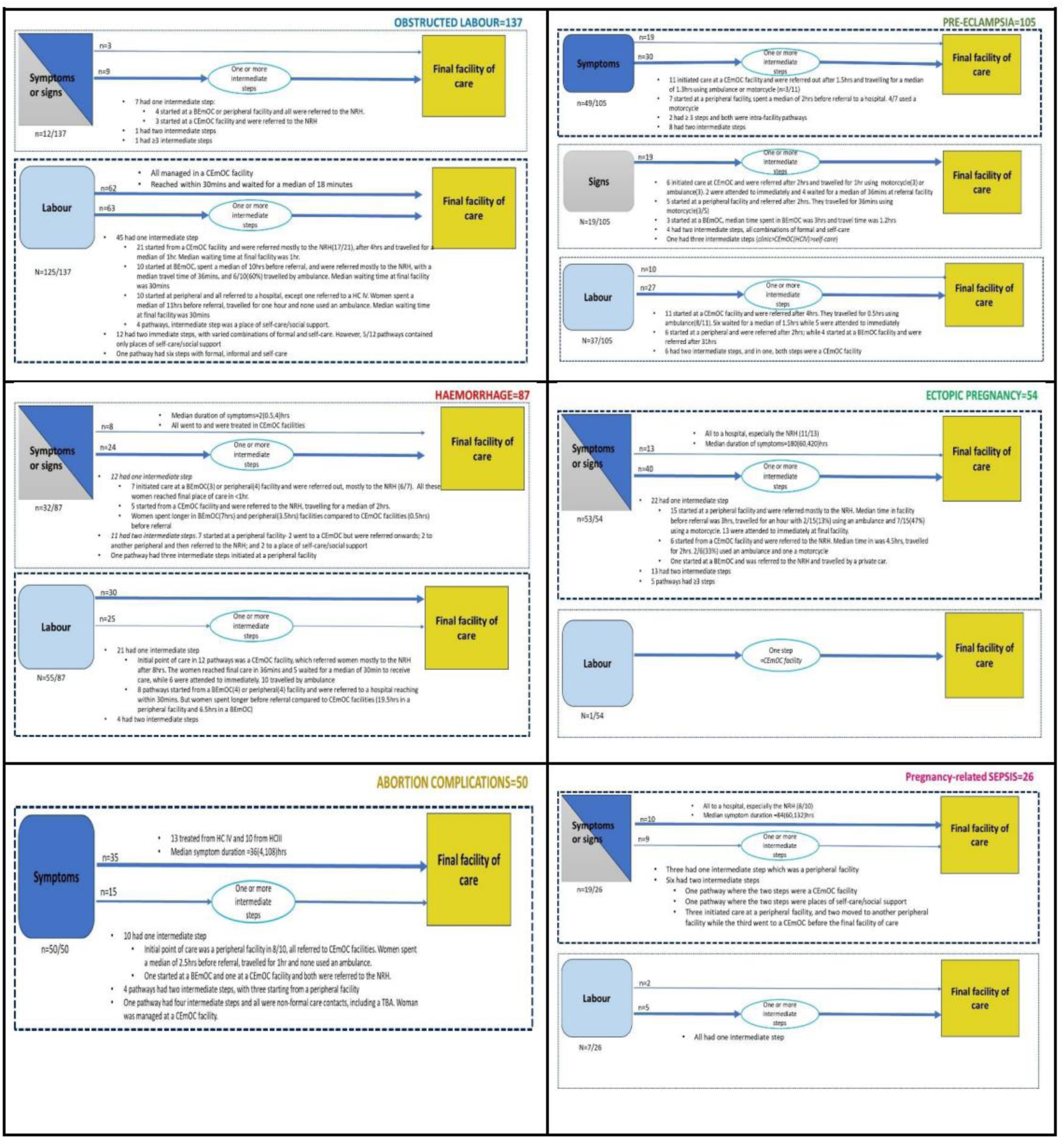
Pathways by obstetric complication (n=433)

**Table 1.** Description of the common pathways to EmOC identified among women who received treatment for obstetric complications in Kamapala (n=433, 9 heath facilities)

| Pathway sequence | Key highlights |
| --- | --- |
| **A=183**<br>*(direct pathways)* | ▪ Care-seeking among women with this pathway started with labour for the majority (55%, 101/183) of whom, 61% (62/101) were treated for obstructed labour.<br>▪ Among the 82 women whose care seeking started with symptoms recognized by the woman, nearly a half were treated for abortion-related complications (43%, 35/82).<br>▪ A majority (53%, 97/183) resided in Kampala, while 43% (79/183) resided in Wakiso and 4% (7/183) lived in other districts.<br>▪ A majority of women travelled on a weekday (81%, 148/183), and 19% (35/183) travelled on a weekend.<br>▪ The most common mode of transport used was the motorcycle (54%, 99/182). Other transport modes were private car (17%, 30/182), public transport (13%, 23/182), special hire (6%, 10/182), ambulance (2%, 4/182) and 3% (5/182) walked.<br>▪ Majority of the women reached the final facility of care within one hour (66%, 118/180), and 34% (62/180) travelled for more than one hour to reach the final facility of care<br>○ The longest travel time was three hours, which was among two cases of ectopic pregnancy complications and one case with sepsis.<br>▪ Among women with this pathway, 25% (46/183) were attended to immediately upon arriving at the facility; while 73% (133/183) waited for a median of 0.3 hours (IQR=0.1-0.8)<br>▪ Among the 183 women, 95 (52%) did not report any challenges with transportation. Among the remaining 88 women, the top three challenges faced with transport were financial constraints (51%, n=45), traffic jam (44%, n=39) and inadequate transport options (31%, n=27). Pot holes were reported by 7% (n=6) of the women.<br>▪ Of all 183 women, 33% (n=60) went to the NRH; 57% (n=105) went to primary and secondary CEmOC facilities; and 10% (n=18)<br>▪ 91% (166/183) of women indicated that they intended to go to the facility where they received care for the complication.<br>○ Reasons for choosing to go to the facility most frequently related to better care (53%, 88/166), convenient access (31%, 51/166) and usual place of care (26%, 43/166). Other reasons given were trust (17%, 28/166), cost (11%, 18/166), being told to go there by other people (15%, 25/166).<br>▪ Among the 17 women who did not intend to go to the facility where they received care, being in critical condition and being brought to the facility by other people was the most common reason given (41%, n=7). Other reasons were being referred by another facility (24%, n=4) and being told/advised by other people (35%, n=6).<br>▪ Instrumental support in form of having someone (partner, relatives, friends/colleagues) to help pay for care or prepare items to go with to the facility was the most reported enabler to care-seeking (93/183, 51%). Other enablers were availability of transport means (21/183, 12%), having prepared items for childbirth (19/183, 10%), and having money (12/183, 7%).<br>▪ Symptoms experienced by the woman were the most reported barrier (25/183, 14%). For example, being in too much pain and travelling on a motorcycle. Having inadequate money to pay for transport was reported by 6% (11/183). |
| **B-pathways with two steps** |  |
| **B1=69**<br>*(referring facility =CEmOC facility)* | ▪ All referrals were made to hospitals, almost all to the tertiary CEmOC hospital (60/69), where they were treated for pre-eclampsia (28/69) and obstructed labor (24/69).<br>▪ Out of the 69, 54% (n=37) travelled for more than one hour to the facility where they received treatment, with majority using an ambulance (34/69, 49%).<br>○ Although, nearly 30% reported moving with a motorcycle (n=19/69).<br>▪ Most (55%, 38/69) spent hours within the CEmOC before being referred with a median duration of 4 (IQR=2,6) hours<br>▪ Overall path had a median duration of 5.6 (IQR=2,36.5) hours.<br>○ The shortest path was 0.6 hours (*referred to a secondary CEmOC and was treated for pre-eclampsia*)<br>○ The longest path was 157 hours (*referred to the tertiary CEmOC and was treated for abortion complications*)<br>▪ Among the 69, 31 reported no challenges with transportation to the final facility of care. Of the remaining 38, 71% (n=27) faced financial constraints, 40% (n=15) reported traffic jam, and 26% (n=10) reported inadequate transport options* as challenges.<br>▪ The most frequently reported enabler for getting help at the CEmOC facility was transport availability (38%, 26/69). Others were having money (n=5), social support (n=11), and being worked on immediately (n=4).<br>▪ Of the 69 women, 37 (54%) did not report any barrier to getting help/care at the CEmOC facility. Less than 10 women reported inadequate money, symptoms and other less frequent (n<5) barriers.<br>Among the 69 women referred directly by a CEmOC facility to the final facility of care |
|  | Women spent a median of 4 hours (IQR=1-33) in the first CEmOC facility visited before being referred. The least time spent was among pathways of women treated for pre-eclampsia (2.3 hours; IQR=0.9-7.5), and the longest time was for the one woman who was treated for abortion complications 156 hours (6.5 days).<br>Women travelled for a median of 1 hour (IQR=0.5-1.5). The shortest travel time was among women treated for hemorrhage (0.7 hours), and longest travel time was among women treated for sepsis (3 hours).<br>51% of women were attended to immediately upon arriving in the final facility of care, and the remaining 49% waited for a median of 1 hour (IQR=0.3-2).<br>Across the six complications, transportation by ambulance was reported by 55% (38/69) of women referred by CEmOC facilities and 32% (22/69) used a motorcycle.<br>Ambulances were used in at least 30% of pathways of women treated for hemorrhage (32%, 12/38) and pre-eclampsia (37%, 14/38); while motorcycle was commonly used by women treated for obstructed labor (50%, 11/22). |
| **B2=66**<br>(referring facility<br>=Peripheral facility) | From the peripheral facility, nearly all women (97%, 63/65) were referred to a hospital, more so to the tertiary CEmOC hospital (40/63).<br>Only two women referred to 1<sup>o</sup> CEmOC.<br>At least 45% travelled for more than 60 minutes to reach the final place of care (n=31/65), with 42% (27/65) using a motorcycle and 22% (14/65) using a private car.<br>A majority of women reported to have spent hours within the peripheral facility prior to being referred, with a median duration of 3 (IQR=1,3) hours.<br>Overall pathway to final care was over 60 minutes in nearly all women (56/65).<br>The shortest pathway was 0.5 hours (*referral to a secondary CEmOC and treated for a ruptured uterus*).<br>The longest path was 87 hours, (*for a woman who spent three days in the peripheral facility before being referred to the tertiary CEmOC hospital where she was treated for ectopic pregnancy complications*).<br>The most common transport challenges faced in reaching final facility of care were traffic jam (53%, 20/38), inadequate transport options (40%, 15/38) and financial constraints (42%, 16/38). Of the 66 women, 28 reported no challenges with transportation.<br>The most frequent enablers to getting care at the peripheral facility were transport availability (20%, 13/66), availability of health workers (17%, 11/66), and having money (15%, 10/66).<br>The most common barriers were the facility lacking capacity to handle the woman's condition (15%, 10/66) and inadequate money (6/66). |
| **B3=29**<br>(referring facility<br>=BEmOC facility) | All women were referred to hospitals, more so to the tertiary CEmOC facility (22/29, 76%), where they were treated mostly for obstructed labor (12/29)<br>Slightly over 50% travelled within 60 minutes, with 12/29 using an ambulance and 10/29 using a motorcycle.<br>A majority (17/29) reported spending hours in the facility before being referred, with a median duration of 5 (IQR=3,7) hours.<br>The median overall path length was 6 (IQR=2.5,14.3) hours<br>The shortest path was 0.6 hours (*referral to a secondary CEmOC, treated for pre-eclampsia*)<br>The longest path was 61 hours (*referral to the tertiary CEmOC, treated for obstructed labor*).<br>Among the 14 women who reported challenges with transportation to the final facility of care, inadequate transport options (n=7, 50%), financial constraints (n=4, 29%) and traffic jam (n=4, 29%) were reported. |
| **C=62**<br>*Pathways with three steps* | The initial point of care was a peripheral facility in 61% (38/62) of pathways.<br>The last point of care before the facility where the complication was treated was a place of self-care or social support in 63% (39/62)<br>32 unique sequences were identified.<br>Activities related to places of self-care/social support included self-medication, getting care or support from family/friend, getting permission to seek care, getting transport/funds/requirements. |
| **D=17**<br>*Pathways with four or more steps* | Observed more in women with OL (7/17)<br>Places of formal care were the first point of contact for 47% (7/17)<br>All pathways were unique to an individual woman<br>Pathways include intra-facility loops in a CEmOC facility (hospital)<br>The longest pathway (by #steps) shows the woman contacted formal providers at peripheral level twice in between informal and self-care.<br>Most of the pathways point to 'community loops' – women moving through/between places of non-formal care. |

\*Includes lack of or unfunctional ambulances.

## Notes

### Competing Interest Statement

The authors have declared no competing interest.

### Funding Statement

This study was funded by the Belgian Directorate for Development Co-operation PhD sandwich programme, awarded to the corresponding author

### Author Declarations

The ethics committees of Makerere University School of Public Health (SPH-2021-169), the Uganda National Council for Science and Technology (HS1952ES), the Institute of Tropical Medicine Antwerp (1529/21) and the University of Antwerp Hospital (2021-1743) gave ethical approval for this work.

